# Integrated diabetes care delivery at primary healthcare facilities in Pakistan: An economic evaluation of a pragmatic randomized controlled trial

**DOI:** 10.64898/2026.10.06.26364849

**Authors:** Nida Khan, Muhammad Amir Khan, Muhammad Ahmar Khan, Shaheer Ellahi Khan

**Author notes:** Correspondence to*: Khan, Nida, Association for Social Development, House No. 12, Street 48, F-7/4, Islamabad, Pakistan., contact number.

## Abstract

**Background:** In Pakistan approximately 7 million people were living with diabetes in 2014. To address the burden of diabetes, a randomized controlled trial was conducted to evaluate the effectiveness of a standardized care package in improving diabetes outcomes in health care settings of Punjab, Pakistan. A costing study was embedded within this RCT to estimate the cost-effectiveness of the standardized diabetes care package in improving diabetes outcomes.

**Methods:** 166 patients were recruited for the costing study from 14 public health clusters in Sargodha district of Pakistan. Analyses was conducted using the societal perspective, broader societal perspective and sensitivity analyses to explore the uncertainty in analyses. Data was analysed for both 166 participants enrolled in costing study and for the 495 participants recruited in the cRCT, using multiple amputations.

**Results:** Using the societal perspective, standardized diabetes care package was found to be cost-effective as compared to the standardized care and was cost saving as the costs in the intervention arm were lower than those in the control arm whereas greater effects were demonstrated in the intervention arm. As per the broader societal perspective and the healthcare perspective only, the standardized care package was not proven to be cost-effective.

## Introduction

In Pakistan approximately 7 million people were living with diabetes in 2014.(1) This figure is estimated to reach about 11.4 million by 2030.(1) Pakistan is ranked 7^th^ in the world for diabetes prevalence and one of the major causes for this ranking is the poor utilization of primary healthcare services. (2,3) According to the Diabetes Prevention Program (DPP) intensive lifestyle and Metformin interventions decrease the incidence of type-2 diabetes. (4) However, compliance with treatment is dependent on various factors which include patient centered factors, therapy-related factors, healthcare system factors, social and economic factors and disease factors. (5,6) Research shows that compliance of patients who have chronic diseases for which treatment could last lifelong, is crucially dependent on the cost of treatment. (7)

Punjab is the largest province in Pakistan, with more than 90 million people living in 36 districts. (8) Planning, financing and delivering health care is the responsibility of the provincial government. At district level, the public infrastructure for delivering health care includes district and tehsil level hospitals and rural health centres. These hospitals and health centres do have doctors and other associated professionals and a laboratory for basic tests such as urine/ blood testing. A district health office is responsible for managing public funded care at these secondary and primary level health facilities. (8)

The province of Punjab has taken lead in taking measures to add integrated diabetes care to these public-funded hospitals and health centers. The Directorate General Health Services Punjab, with the support of technical and development partners, undertook an intervention development and evaluation activity for delivering integrated diabetes care at primary and secondary level facilities. The intervention package developed, through DGHS-led technical working group, included: case management desk-guide, material to train doctors and allied staff, and pictorial tool to counsel on lifestyle change.

The primary aim of this health economic analysis is to assess the cost-effectiveness of an improved quality package of care for patients with diabetes in rural Pakistan, compared to standard care. The analysis will consist of a within trial analysis of the costs and effects associated with each arm of the trial.

## Methods Overview

The methods used to conduct this analysis were guided by the WHO reference case for economic evaluations of healthcare (9). A societal perspective was adopted for the analysis. The cost and benefits were calculated for the study period of nine months. No discounting was required as the time horizon was less than one year. The primary analysis consisted of a comparison of the cost-effectiveness of integrated delivery of routine care for diabetes compared to standardised package of care in Pakistan and was conducted for the patients who completed the costing questionnaire. These patients represent the complete cases and are a subsample of 166 patients out of the full dataset. We also conducted a secondary analysis of the same comparison to include all patients so that all evidence on costs and effects could be combined in the analysis. Additionally, we conducted a costing analysis to assess the costs associated with implementing the care package used in the intervention arm of the trial.

### Trial design and data collection

Our economic analysis was conducted alongside a pragmatic cluster randomised controlled trial which compared a standardised package of integrated care for diabetes (intervention arm) versus standard care practices (control arm) at primary health facilities (10). The trial aimed at assessing whether the adoption and implementation of this intervention package in the primary healthcare setting resulted in better diabetes control. The trial was conducted in 14 public health facilities in Sargodha district and was given ethical approval by The University of Leeds Research Ethics Committee (reference number: HSLTLM11019), the National Bioethics Committee (NBC) Pakistan (reference number: NBC: 90) and has been registered with the Current Controlled Trials (Trial registration number: ISRCTN23905886). Randomisation was conducted at the facility level due to the training and extra resources required in the intervention group. Health facilities were randomised in a 1:1 ratio to each of the treatment groups. The facilities were randomised to give usual care or standardised care package which included recommended drug prescription according to international guidelines and lifestyle modification counselling. In addition to this, the intervention arm patients received calls to remind them about their follow up one to two days before it was due. The trial registered a total of 495 patients, 250 in intervention and 245 in control arm. Thirty-one patients were lost to follow up. This included 10 deaths and 21 patients who withdrew from the trial. The participants in the trial were newly diagnosed diabetes patients aged twenty-five years and above. The patients were followed for a period of 9 months and their glucose levels, other clinical outcomes and medicines prescribed were recorded in a non-communicable disease (NCD) card.

### Health Outcomes

The primary outcome of interest for the cost-effectiveness analysis is the incremental reduction of HbA1c score. HbA1c score was recorded for each patient at baseline and 9 months later at end-line on their NCD card. The change in HbA1c from baseline to end-line was then calculated for each patient. For the incremental analysis we consider the reduction of HbA1c of 0.5 as the meaningful unit of health benefit so that incremental cost-effectiveness ratios are presented as the cost per incremental reduction of 0.5 in HbA1c score.

### Resource use and costs

The trial NCD cards were completed by healthcare facility staff on behalf of patients at each visit to the healthcare facility, and recorded the number of healthcare visits each patient had, any equipment that was used, tests that were performed and medications prescribed. This resource use information was supplemented by information from a patient questionnaire. This questionnaire was completed by a sub sample of 166 patients of those registered in the trial. This questionnaire captured both direct costs, such as fees for services and additional costs for medicines, and indirect costs, such as travel costs and loss of earnings, incurred by patients. This questionnaire also collected information on the regular costs patients incurred due to lifestyle modification such as due to necessary changes in diet, physical activity and smoking.

We converted all resource use data to costs using appropriate unit costs estimated at the time of analysis. Unit costs are reported in **Table 1**. We assigned costs on a per unit basis with unit values based on data collected within the trial such as staff costs, medication costs and unit costs for equipment.

**Table 1:** Unit costs.

| Type of service | Cost PKR (USD\$) | Unit of measure | Notes | Source |
| --- | --- | --- | --- | --- |
| Doctor (registration visit/<br>follow up visit) | 38.45 (0.36) | Per visit | Doctor, per patient contact<br>lasting 5 minutes | Trial data |
| Paramedic (registration<br>visit) | 48.15 (0.45) | Per registration visit | Paramedic, per patient<br>contact lasting 15 minutes |  |
| Paramedic (follow up visit) | 16.05 (0.15) | Per follow up visit | Paramedic, per patient<br>contact lasting 5 minutes |  |
| NCD card | 4.7 (0.044) | Per card |  |  |
| Brochure | 10.6 (0.100) | Per brochure |  |  |
| Needles | 6.1 (0.058) | Per visit |  |  |
| Test strip | 27 (0.25) | Per visit |  |  |
| Urine strip | 7.8 (0.074) | Per visit |  |  |
| Referral slip | 3.2 (0.030) | Per referral |  |  |
| **Implementation costs** |  |  |  |  |
| Type of service | Cost PKR (USD\$) | Unit of measure | Source | |
| Training for doctors | 4000 (38.09) | Per doctor trained | Trial data |  |
| Training for paramedics | 4000 (38.09) | Per paramedic<br>trained |  |  |
| Doctor's manual | 418 (3.98) | Per manual |  |  |
| Paramedic's manual | 274 (2.60) | Per manual |  |  |
| Flipbook | 570 (5.42) | Per flipbook |  |  |
| Desk guide | 274 (2.60) | Per desk guide |  |  |
| **Medication** | **Dose** | **Cost per dose**<br>**PKR (USD\$)** | **Unit cost PKR (USD\$)** | **Description** |
| Neodipar | 1500mg* | 4.32 (0.04) | 1.44 (0.013) | Unit cost per tablet, dose is daily dose, and cost per dose is cost per day. |
| Amaryl | 1mg* | 7.52 (0.071) | 7.52 (0.071) | Per tablet, 1 per day |

We assumed that costs of trial medication fell to the healthcare provider but the cost of any other medications were captured in the medication costs reported by patients. We calculated per patient cost for each patient as the sum of the cost of all health visits. We calculated the unit cost of a health visit from a healthcare provider perspective by multiplying the duration of each health visit by the costs of staff time of the health professional. This provided us the cost to the health facility for each time a patient visit. As the duration of visits to the health facility and the type of health professional who took the appointment was not recorded, we made an assumption about the average length of time of a visit to the health facility and what level of health professional is seen. These assumptions were based on discussion with the researchers on the ground who conduct the trial and have some knowledge about how long the visits last on average and who takes the appointments. Total costs to the healthcare provider were calculated for the intention to treat (ITT) population. We generated the summary by intervention group and differences in costs between groups were compared using independent sample t-tests.

### Implementation Cost

We conducted a cost analysis to assess the implementation costs associated with the intervention arm. These costs represent the investment cost needed to enable the quality of care provided by the intervention arm and are relevant if the intervention is to be adopted and scaled-up nationally. However, as an upfront cost they do not reflect a per-patient cost. We present these costs as the cost that will be incurred for each new facility to adopt the quality of care used in the intervention arm of the trial. We also analysed the costs to the healthcare provider at the point of treating patients.

### Statistical Analysis

#### Primary analysis

In the cost-effectiveness analysis we adopted an intention-to-treat (ITT) perspective for analysing and summarising the health economic trial data. For the primary analysis, we limited the ITT population to those patients who completed the costing questionnaire as well as the other trial NCD card. We calculated the mean costs and outcomes from the observed data over the trial period of 9 months. We calculated incremental cost-effectiveness ratios to show the cost per incremental reduction in HBA1C score of 0.5. As health benefit is measured in natural units there is no pre-defined cost-effectiveness threshold value of willingness to pay per incremental reduction in HBA1C score. Therefore, we present the cost-effectiveness results to enable decision makers to make informed decisions based on the relevant willingness to pay threshold in their context.

In adherence to the WHO reference case, we used a societal perspective for costs in the base case. This included per patient cost to healthcare providers and out of pocket patient cost for healthcare provider (i.e., costs that displace costs that would otherwise fall on a healthcare budget e.g., medication costs).

To explore the uncertainty in the primary analysis we conducted the three sets of scenario sensitivity analysis. Firstly, we analysed using a broader social perspective in which we included indirect patient costs i.e., per patient costs to healthcare providers and the out-of-pocket costs for healthcare provider provided plus indirect costs e.g., travel and time. Secondly, we conducted a sensitivity analysis from the healthcare provider perspective which included per patient costs incurred by healthcare providers but excluded all out-of-pocket patient costs. Thirdly, we explored the effect of adjusting for baseline differences in HbA1c score.

#### Secondary analysis

The primary analysis was followed by a secondary analysis in which we ran all of the above analysis using the full dataset of patients and imputing cost data for those that did not complete the patient costing questionnaire. This enabled a cost-effectiveness analysis in which we were able to include all of the data on costs and outcomes that was collected in the trial.

### Missing data

Seven patients had missing end line HbA1c scores. We imputed the missing data using the multiple imputation approach which is the recommended method of imputation for economic evaluation alongside clinical trials as it includes randomness to reflect the uncertainty inherent in missing data by using iterative multivariable regression techniques (11). The patient cost data was collected for only 166 of the 495 patients. For the secondary analysis, to enable an analysis of the full ITT population, we used mean imputation methods by trial arm to impute the missing patient cost data.

## Results

### Resource use and costs

We converted the healthcare resource to healthcare costs using the relevant unit costs. The costs to the healthcare provider are presented for both the complete case population used in the primary analysis and the ITT sample used in the secondary analysis but the patient costs are only presented for the 166 (complete cases) patients who completed the costing questionnaire. The analysis of the difference in healthcare costs from a societal perspective found that there was no significant difference between the intervention and control arms. However, analysis of the difference in costs from a broader societal perspective indicated there was a statistically significant difference in costs (p<0.01) when patient indirect costs were included. This shows the importance of increased out of pocket costs to patients. Incremental lifestyle modifications costs were reported to be non-existent by the participants in intervention arm.

### Implementation costs

We found the cost of implementing the intervention in a healthcare facility to be 19,072 PKR per facility. That is, if this intervention was to be implemented in more healthcare facilities, this is the upfront cost per facility to put this intervention in place. However, if the intervention was to be implemented nationally, there could be additional cost savings through economies of scale.

### Health outcomes

Measurements of HbA1c at baseline and end line are shown in **Table 2** along with the change in HbA1c score over the trial period, for both the complete case population used in the primary analysis and the ITT sample used in the secondary analysis. There was a reduction in HbA1c scores over the period of nine months in both treatment arms, but a greater reduction is observed in the intervention arm. Statistical tests indicated there was no significant difference in HbA1c score at baseline but we explored this further in a sensitivity analysis by adjusting for baseline differences. Independent sample t-tests indicated that in the ITT sample there was a statistically significant difference (p<0.01) between treatment groups in the change in hba1c score. The same test for the complete case sample showed no significant difference but this is likely due to being under-powered in the smaller sample.

**Table 2:** Base case results – Costing patients only, disaggregated societal perspective.

|  | Intervention arm *n*=83 |  | Control arm *n*=83 |  |
| --- | --- | --- | --- | --- |
| **Time point** | **Mean** | **SD** | **Mean** | **SD** |
| Baseline HBA1C | 10.077 | 2.446 | 10.034 | 2.612 |
| Endline HBA1C | 8.243 | 2.501 | 8.765 | 2.6 |
| Change in HBA1C | -1.835 | 3.285 | -1.269 | 2.911 |

### Cost-effectiveness and decision uncertainty

#### Primary analysis

The cost effectiveness results from the primary analysis are shown in **Table 3**. The treatments have been arranged in order of cost so that the incremental costs and effects refer to a comparison of neighbouring treatments in the table.

**Table 3:** Primary Analysis: Patients with cost data.

| Treatment group | Mean ± SD<br>Cost PKR (USD\$) | Incremental cost PKR (USD\$) | Change in HBA1c | Incremental change | ICER per 0.5 reduction in HBA1C PKR (USD\$) |
| --- | --- | --- | --- | --- | --- |
| ***Societal perspective*** |  |  |  |  |  |
| Intervention | 15,278.38 ± 6220.39 (145.50 ± 59.23) | | -1.835 (3.285) | | |
| Control | 15,632.46 ± 5058.32 (148.87 ± 48.17) | 354.08 (3.37) | -1.269 (2.911) | -0.566 | dominated |
| ***Broader societal perspective*** |  |  |  |  |  |
| Control | 29,649.21 ± 10961.64 (282.37 ± 104.39) | | -1.269 (2.911) | | |
| Intervention | 34,475.2 ± 22954.5 (328.33 ± 218.60) | 4825.99 (45.96) | -1.835 (3.285) | 0.566 | 4,263.24 (40.6) |
| **Sensitivity Analysis: Healthcare provider costs only** |  |  |  |  |  |
| Control | 2,374.37 ± 1026.94 (22.60 ± 9.77) | | -1.269 (2.911) | | |
| Intervention | 2,809.96 ± 883.67 (26.75 ± 8.41) | 435.59 (4.14) | -1.835 (3.285) | 0.566 | 348.80 (3.31) |
| **Societal Perspective – adjust for baseline differences in HBA1C score** |  |  |  |  |  |
| Intervention | 15,278.38 ± 6220.39 (145.50 ± 59.23) | | -1.819 (0.22) | | |
| Control | 15,632.46 ± 5058.32 (148.87 ± 48.17) | 354.08 (3.37) | -1.285 (0.211) | -0.534 | dominated |

The results from the societal perspective (the base case) in the primary analysis show that the intervention arm is more cost-effective than the control arm and could be cost saving as the costs in the intervention arm are lower than those in the control arm whereas greater effects are achieved in the intervention arm. That is, the control arm of the trial is dominated by the intervention arm due to the cost saving and greater health gains.

#### Secondary analysis

The cost-effectiveness results from the secondary analysis are shown in **Table 4**. The results of the secondary analysis using the full dataset and imputing missing patient cost data found the result from the societal perspective (base case) in the primary analysis to be robust. This analysis shows even greater benefit over the whole trial population and still shows lower costs associated with the intervention arm compared to the control arm.

**Table 4:** Secondary Analysis: All patients – *Societal Perspective (mean imputation of patient costs)*

| Treatment group | Cost (PKR) | Incremental cost | Change in HBA1c | Incremental change | ICER (cost per 0.5 reduction in HBA1C) |
| --- | --- | --- | --- | --- | --- |
| Intervention *n*=240 | 13,994.91 ± 3768.82 (133.27 ± 35.88) |  | -2.2 (3.197) |  |  |
| Control *n*=224 | 14,517.18 ± 3192.01 (138.25 ± 30.4) | 522.27 (4.97) | -1.384 (2.878) | -0.816 | dominated |
| *Broader Societal Perspective* |  |  |  |  |  |
| Control | 28,533.93 ± 6705.51 (271.74 ± 63.85) |  | -1.384 (2.878) |  |  |
| Intervention | 33,391.68 ± 13479.95 (318 ± 128.37) | 4857.75 (46.25) | 2.2 (3.197) | 0.816 | 2976.56 (28.34) |
| Sensitivity analysis: Healthcare provider costs only |  |  |  |  |  |
| Control *n*=224 | 1,259.09 ± 1080.84 (11.99 ± 10.28) |  | -1.384 (2.878) |  |  |
| Intervention *n*=240 | 1,526.44 ± 1093.81 (14.53 ± 10.40) | 267.35 (2.54) | 2.2 (3.197) | 0.816 | 163.82 (1.55) |
| Societal Perspective – adjust for baseline differences in HBA1C score |  |  |  |  |  |
| Intervention | 13,994.91 ± 3768.82 (133.27 ± 35.88) |  | -2.073 (0.205) |  |  |
| Control | 14,517.18 ± 3192.01 (138.25 ± 30.04) | 522.27 (4.97) | -1.519 (0.238) | -0.554 | dominated |

### Sensitivity analysis

We conducted three sensitivity analyses to test the robustness of results in both primary and secondary analysis. The sensitivity analyses included analysis of: a) a broader societal perspective on costs, b) healthcare provider costs only, and c) adjusting for baseline differences in HbA1c score. The results of each sensitivity analysis are presented in **Table 3** and **Table 4** for primary and secondary analysis, respectively. The results of the sensitivity analyses in which adjustment are made for baseline differences in HBA1C score, find the base case results to be robust in both the primary and secondary analyses. When taking a broader societal perspective on costs, the results of the cost-effectiveness analysis do not show a cost-saving and this is driven by the additional costs incurred by patients for travel to the healthcare centre for additional healthcare visits in the intervention arm. In this case the ICER shows that the cost per incremental reduction in HBa1c of 0.5 is 4,263.24 PKR (USD$ 40.6) in the primary analysis and 2,976.56 PKR (USD$ 28.34) in the secondary analysis. Similarly, in the analysis of healthcare provider costs only, the results do not show a cost saving but the difference here is minimal given the health benefits. From the healthcare provider perspective only, the ICER shows that cost per incremental reduction in HBA1C of 0.5 is 348.80 PKR (USD$ 3.3) in the primary analysis and 163.82 PKR (USD$ 1.5) in the secondary analysis. The lower ICERs in the secondary analysis are a result of the higher average gains when all of the clinical data is included in the analysis.

## Discussion

We found the diabetes care management package to be cost effective as its health gains were greater as compared to routine care being provided at public health facilities. Primary analysis finds that the interventions is rather cost-saving as it produced larger health gains for a reduced cost compared to control. The healthcare costs for additional reduction in HbA1c were found to be minimal in all perspectives. However, depending on the cost perspective adopted, the differences in costs vary. Using the societal perspective, where only direct cost for patients and providers were taken into account, the diabetes care package was found to be cost-effective in reducing 0.5 HbA1c as compared to control arm. Economic evaluations of similar diabetes care packages with Samoan (12) and Black American populations (13) have also been found to be cost-effective.

We found that our diabetes care intervention package costs less than the routine care provided for diabetes and is more effective. Less cost of the intervention could be attributed to the integration of diabetes care package within the already existing public healthcare centres. A recent systematic review showed that the integrated care models for diabetes patients might be more cost-effective (14). Intervention was shown to be cost-effective for both the subsample included in the costing study and for the complete study sample recruited in the diabetes cRCT. Accounting for the indirect medical costs incurred by patient using the broader societal perspective, the diabetes management intervention was not found to be cost-effective which speaks to the high travel costs incurred by the patients for visiting healthcare facilities for accessing diabetes care. It is important to note that these cost-effectiveness results may be underestimated as this analysis is based on only one measure of health benefit: reduction in HbA1c score over a small trial period, a limitation that has also been reported by Zhang et al (15). This study may also be underestimating the cost saving as it does not take into account the other social factors and future costs that could be incurred on related comorbid conditions by both the patients as reported by Lin. et al (16). Costs incurred on treatment of diabetes, in addition to the co-morbidity related costs could be catastrophic in the long term (17).

The data shows that the lifestyle modification component in the intervention arm does not imply any extra cost to the patient as compared to control arm in order to manage their diabetes. Lifestyle modification component did not imply any additional cost in the intervention arm as compared to the control arm. However, many other studies have reported significant costs associated with lifestyle modifications recommended for Diabetes control (18). The minimal cost of modifications in lifestyle in this intervention could be attributed to the intervention being contextualized for the population. The suggested modifications were integrated into their existing dietary practice. For example, the suggestion to take out a small portion of the regular meal before adding extra oil, salt and/ or sugar, was a modification that did not imply any special diet plans. Therefore, the study shows that not only scaling this intervention to other similar areas in Pakistan could help in controlling diabetes in the country, but also other low-resource countries could adapt these modifications according to their local context.

One of the major strengths of this study is that it was conducted alongside a pragmatic randomized controlled trial. The exclusion criteria were also minimal which makes the study generalizable on patients with profiles similar to those that were included in this trial. Additionally, conducting analysis from a societal perspective and presenting results in a disaggregated format means that the results presented are relevant for policy makers in countries with a range of different healthcare systems. The intervention tested had minimal impact on existing clinical work flows and owing to the pragmatic design, allowed tailoring to site-specific requirements.

The main weakness of the study was the short period of follow up (i.e., 9 months), as dictated by the funding constraints. This period may have been too short to observe all the effects and costs of the standardised care package. This is particularly important for diabetes as there may be many long-term benefits through improved management of diabetes such as avoidance of co-morbidities. However, despite this short follow up period we did observe changes in the HbA1c levels. Also, there could be chances of recall bias for the patient questionnaire as 9 months is a long period.

## Conclusion

We found the use of a standardised care package for Diabetes at public healthcare centres to be cost-effective, as compared to the routine care. The cost-effectiveness of the diabetes care package implies that there is an economic justification for public sector investment in scaling the intervention.

## Data Availability

All data produced in the present study are available upon reasonable request to the authors

## Acknowledgements

We acknowledge the support of Farooq Manzoor, Provincial Manager, Non-Communicable Disease Control Program, Lahore, Pakistan.

